# Excimer LASER coronary atherectomy for ST-segment elevation myocardial infarction: a multicenter retrospective cohort analysis

**DOI:** 10.1101/2024.05.30.24308248

**Authors:** Yuji Matsuda, Taishi Yonetsu, Ken Kurihara, Shigeo Shimizu, Akihiko Matsumura, Hiroshi Inagaki, Yuko Onishi, Kaoru Sakurai, Takaaki Tsuchiyama, Takashi Ashikaga, Hiroyuki Fujii, Kazuo Kobayashi, Ikhtiyorjon Khamdamov, Yosuke Yamakami, Tomoyo Sugiyama, Tomoyuki Umemoto, Tsunekazu Kakuta, Tetsuo Sasano

**Affiliations:** Department of Cardiovascular Medicine, Tokyo Medical and Dental University, Tokyo, Japan; Department of Cardiovascular Medicine, Ome Medical Center, Tokyo, Japan; Department of Cardiology, National Hospital Organization Disaster Medical Center, Tokyo, Japan; Department of Cardiology, Kameda Medical Center, Chiba, Japan; Department of Cardiovascular Medicine, Soka Municipal Hospital, Saitama, Japan; Department of Cardiology, Hiratsuka Kyosai Hospital, Kanagawa, Japan; Department of Cardiovascular Medicine, Shin-Yurigaoka General Hospital, Kanagawa, Japan; Department of Cardiology, Tokyo Metropolitan Hiroo Hospital, Tokyo, Japan; Department of Cardiology, Musashino Red Cross Hospital, Tokyo, Japan; Department of Cardiology, Yokohama Minami Kyosai Hospital, Kanagawa, Japan; Department of Cardiology, Kashiwa Municipal Hospital, Chiba, Japan; Department of Cardiovascular Medicine, Graduate School of General Medical and Dental Science, Tokyo Medical and Dental University, Tokyo, Japan; Division of Cardiovascular Medicine, Tsuchiura Kyodo General Hospital, Ibaraki, Japan

**Keywords:** ST-segment elevation myocardial infarction, excimer laser coronary atherectomy, major adverse cardiovascular events, percutaneous coronary intervention

## Abstract

**BACKGROUND:** Excimer laser coronary atherectomy (ELCA) is used for thrombotic culprit lesions in ST-segment elevation myocardial infarction (STEMI), but its efficacy is still unclear. The study objective was to investigate the clinical outcomes of STEMI patients after ELCA.

**METHODS:** Data of consecutive patients undergoing primary percutaneous coronary intervention (PCI) within 24 hours of onset, in 12 healthcare facilities in Japan, were retrospectively analyzed. Patients were divided into ELCA and non-ELCA groups. The primary endpoint was target vessel-related major adverse cardiac events (TV-MACE). Cox regression analysis and propensity score matching were performed to adjust for selection bias in the cohort.

**RESULTS:** A total of 2593 patients, which included 427 patients treated with ELCA, were analyzed with a median follow-up of 815 (390–1385) days. There was no significant difference between the two groups in terms of TV-MACE-free survival rate. ELCA use was not a significant determinant of TV-MACE (hazard ratio [HR] 1.265, 95% confidence interval [CI], 0.910–1.757; p=0.161). Nevertheless, when the ELCA group was stratified by the ELCA catheter size, the large catheter (1.4 mm–1.7 mm) group showed better clinical outcomes than the others in univariate Cox regression analysis (HR 0.30, 95% CI 0.10–0.95, p=0.040). In the propensity score-matched cohort of 736 patients (368 pairs), the TV-MACE-free survival did not differ between the two groups.

**CONCLUSIONS:** ELCA did not show clinical benefit in terms of the rate of adverse cardiac events in patients with STEMI. There was evidence of efficacy when a large ELCA catheter was used, warranting further prospective studies.

**Clinical Perspective:** *What is new?:*

- In a relatively large-scale registry of STEMI patients undergoing primary PCI, which included 427 patients treated with ELCA, the use of ELCA did not show clinical benefits in reducing target-vessel related adverse events.
- The use of ELCA was not associated with improved coronary flow or myocardial perfusion, but rather with higher peak values of cardiac markers.
- Larger ELCA catheters (≥1.4mm diameter) may be associated with better clinical outcomes compared to smaller (0.9mm) ELCA catheters, suggesting potential areas for future research.

*What are the clinical implications?:*

- The routine use of ELCA may not reduce adverse cardiac events in primary PCI for patients with STEMI.
- The use of ELCA should be limited to lesions where large-sized ELCA catheters can be safely applied.

## Introduction

Acute myocardial infarction (AMI) typically arises from thrombus formation in the coronary arteries due to the disruption of lipid-containing atherosclerotic plaques. ST-segment elevation AMI (STEMI) is a subtype of AMI in which immediate revascularization via percutaneous coronary intervention (PCI) shows significant clinical benefits over drug-based treatments, including thrombolysis. Widespread application of primary PCI for patients with STEMI, combined with advances in periprocedural pharmacotherapy and procedural technology, including effective antithrombotic agents and drug-eluting stent implantation, has led to reduced in-hospital mortality.^1,2^ However, the rate of decline in in-hospital mortality in patients with STEMI has slowed.^3^ One of the challenges in primary PCI for patients with STEMI is the minimization of massive embolization of thrombus and atherosclerotic plaque components. Although multiple devices, such as thrombectomy catheters and distal protection devices, have been introduced, none have proven significant clinical benefits in routine use. Excimer light amplification by stimulated emission radiation (LASER) coronary atherectomy (ELCA) has been used in PCI since its introduction in the 1990s.^4^ Light is absorbed by the plaque and thrombus, which releases a pressure wave that initiates rapid fluid displacement, leading to the vaporization and debulking of plaques.^5^ Furthermore, ELCA vaporizes thrombus^6^ and suppresses platelet aggregation.^7^ ELCA has been utilized in myriad PCI settings, including acute coronary syndrome (ACS) resulting from thrombotic lesions.^8^ STEMI is thought to receive the greatest benefit from ELCA, given its large thrombus and plaque burden. Nevertheless, the effectiveness of ELCA in PCI is still controversial, given the limited number of studies and lack of large randomized control trials. Some small-sized, single-center studies have reported better myocardial reperfusion and a lower rate of adverse cardiac events in patients with STEMI treated with ELCA than in those treated without ELCA,^9,10^ whereas other studies failed to show improved outcomes with ELCA.^11^ Previous research may not have accurately assessed clinical outcomes post-ELCA because of the significant selection bias in favor of ELCA use. To address this, we conducted a comprehensive, real-world, retrospective cohort study, including centers where ELCA is a common practice in primary PCI and others where it is not available.

## Methods

### Study population

The ST-Elevation myocardial infarction treated with Laser Atherectomy (STELA) registry (UMIN-CTR 000043818) is a retrospectively collected, multicenter registry of patients with STEMI who underwent primary PCI within 24 h from the onset of myocardial infarction. Twelve regional healthcare centers in Japan providing continuous emergency care, including primary PCI for STEMI, participated in this study. ELCA was available at seven of the participating sites. Consecutive patients with STEMI who underwent primary PCI at the participating sites within 24 h of symptom onset between January 2015 and December 2019 were enrolled. The exclusion criteria were refusal or waiver of agreement to participate, loss of follow-up data at 30 d after primary PCI, and multiple culprit lesions identified at baseline.

### Ethical approval

The study protocol was approved by the Institutional Ethics Committee of each participating hospital. This study was performed in accordance with the principles of the Declaration of Helsinki.

### Clinical data collection

Data, including baseline clinical characteristics, laboratory data, medications at discharge, procedural information, information on outpatient visits, and subsequent adverse events, were retrospectively collected from the medical records at each participating site. Cardiac enzymes were measured every 3-6 h according to the discretion of the attending physicians until the cardiac enzymes decreased; the maximum values were designated the peak values. Coronary angiograms were digitally recorded, de-identified, and sent to the imaging laboratory at Tokyo Medical and Dental University for image analysis.

### Primary PCI procedures

Decisions regarding initial management in the emergency department depended on the attending physicians, and the procedures during primary PCI depended on the operator’s discretion, including the use of ELCA in primary PCI. In ELCA-available hospitals, ELCA was performed using a pulsed-wave xenon chloride excimer laser (X-80 Vitesse RX, Phillips, Amsterdam, The Netherlands) at a wavelength of 308 nm, pulse duration of 135 ns, and output of 165 mJ/pulse. The catheter size (0.9, 1.4, or 1.7 mm), fluence (30–80 mJ/mm^2^), and pulse repetition rate (25–80 Hz) were selected at the operator’s discretion. Patients were treated pharmacologically and with interventional procedures that complied with contemporary guidelines.^12^

### Angiographic analysis

Angiographic data were analyzed by independent investigators who were blinded to the patients’ demographic and outcome data. Quantitative coronary angiography (QCA) analyses were performed on the baseline and final angiograms for each patient using a dedicated software (QAngio, Medis, The Netherlands). The TIMI flow grade was determined at baseline and on final angiography. The corrected TIMI frame count was evaluated^13^ adjusting by the frame rate to 30 fps in the final angiograms. The TIMI thrombus grade was assessed at baseline.^14^

### Endpoints

The primary endpoint was target vessel-related major adverse cardiac events (TV-MACE), defined as a composite of cardiovascular death (CVD), target vessel revascularization (TVR), and target vessel non-fatal myocardial infarction (TV-MI). The secondary endpoints were each component of TV-MACE, peak values of cardiac enzymes, angiographical results of primary PCI, including the final TIMI flow grade, and corrected TIMI frame count.

### Statistical analysis

Patients were divided into two groups regarding the use of ELCA during the primary PCI (ELCA and non-ELCA), and the endpoints were compared between the two groups. Statistical analyses were performed using R version 4.3.2 (R Foundation for Statistical Computing, Vienna, Austria). Categorical variables were expressed as numbers with percentages and were compared using the chi-square test. Continuous valuables were expressed as the mean ± standard deviation when normally distributed, or median with inter-quartile range for non-parametric variables. Continuous variables were compared using the t-test or Mann–Whitney U test, as appropriate. Statistical significance was set at p < 0.05. Multivariate logistic regression analyses were performed to determine the predictors of periprocedural clinical outcomes. For statistical time-dependent comparisons, the log-rank test and Cox proportional hazards analysis were performed. Stepwise regression guided by the Akaike information criterion was used to fit the regression model. The propensity score (PS) was derived for each patient based on variables that showed statistically significant differences between the ELCA and non-ELCA groups, and clinically relevant variables at baseline. PS matching (PSM) was then performed with a 1:1 algorithm using nearest-neighbor matching with a caliper width of 0.2, standard deviation of the logit of the PS, and no replacement, which yielded 368 matched pairs. The primary and secondary endpoints were compared between the two matched groups.

## Results

Regarding study population of the total cohort, from a total of 2777 patients enrolled in the registry, 184 patients were excluded from the analysis: 3 patients with insufficient angiographic data, 134 patients lost to follow-up within 30 days, 40 patients with multiple culprit lesions treated in the primary PCI, and 7 patients with bypass graft failure. The final dataset comprised 2593 patients including 427 (16.5%) patients who underwent ELCA and 2166 (84.6%) patients who did not undergo ELCA (Figure 1 and Table 1).

**Figure 1.**
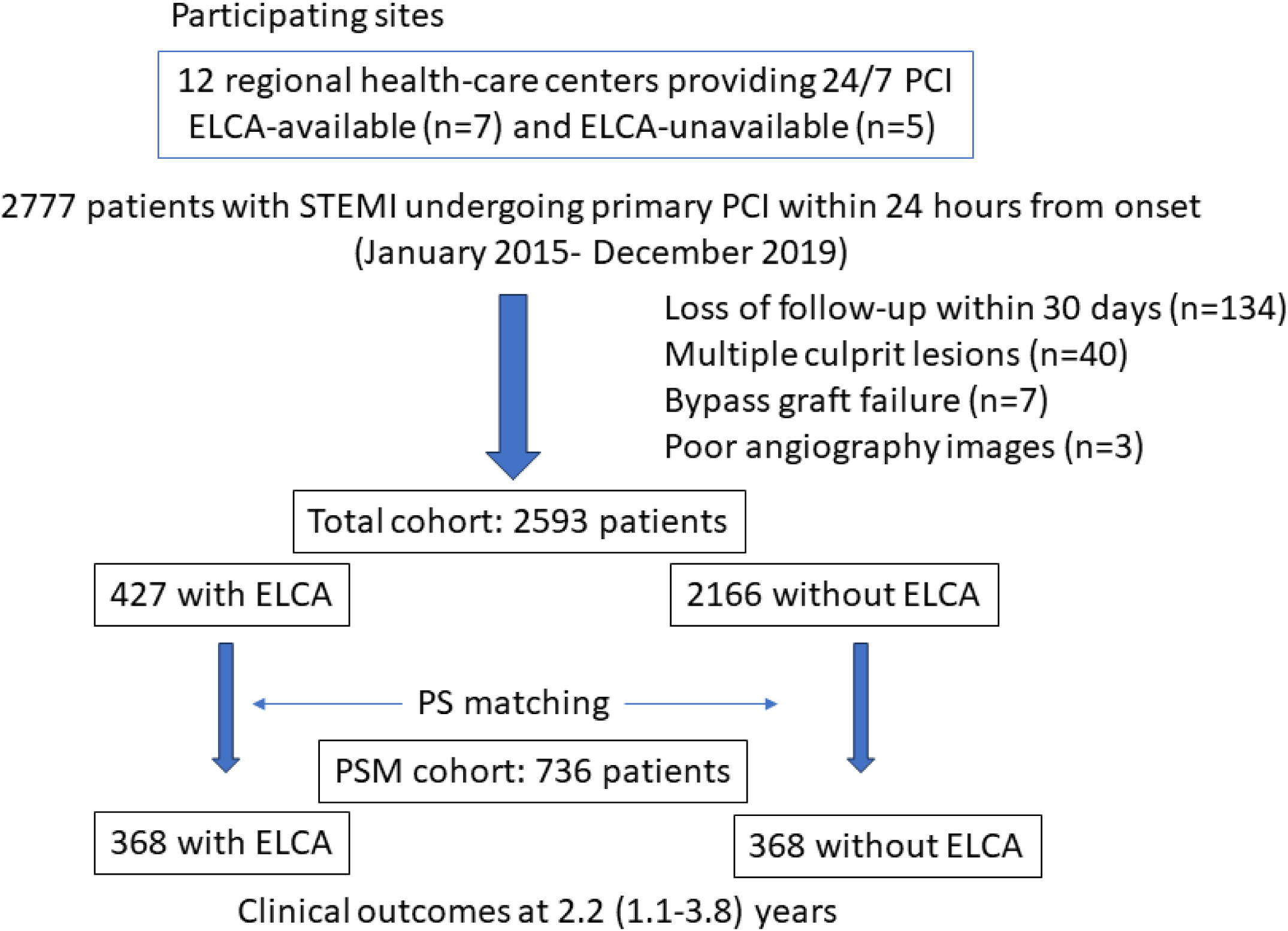
Patient population. Study population from the STELA registry. A total of 2777 patients from 12 hospitals in Japan were enrolled. The study cohort comprised 2593 patients, including 427 patients treated with ELCA. ELCA, excimer laser coronary atherectomy; PS, propensity score; PSM, propensity score matching.

**Table 1.**
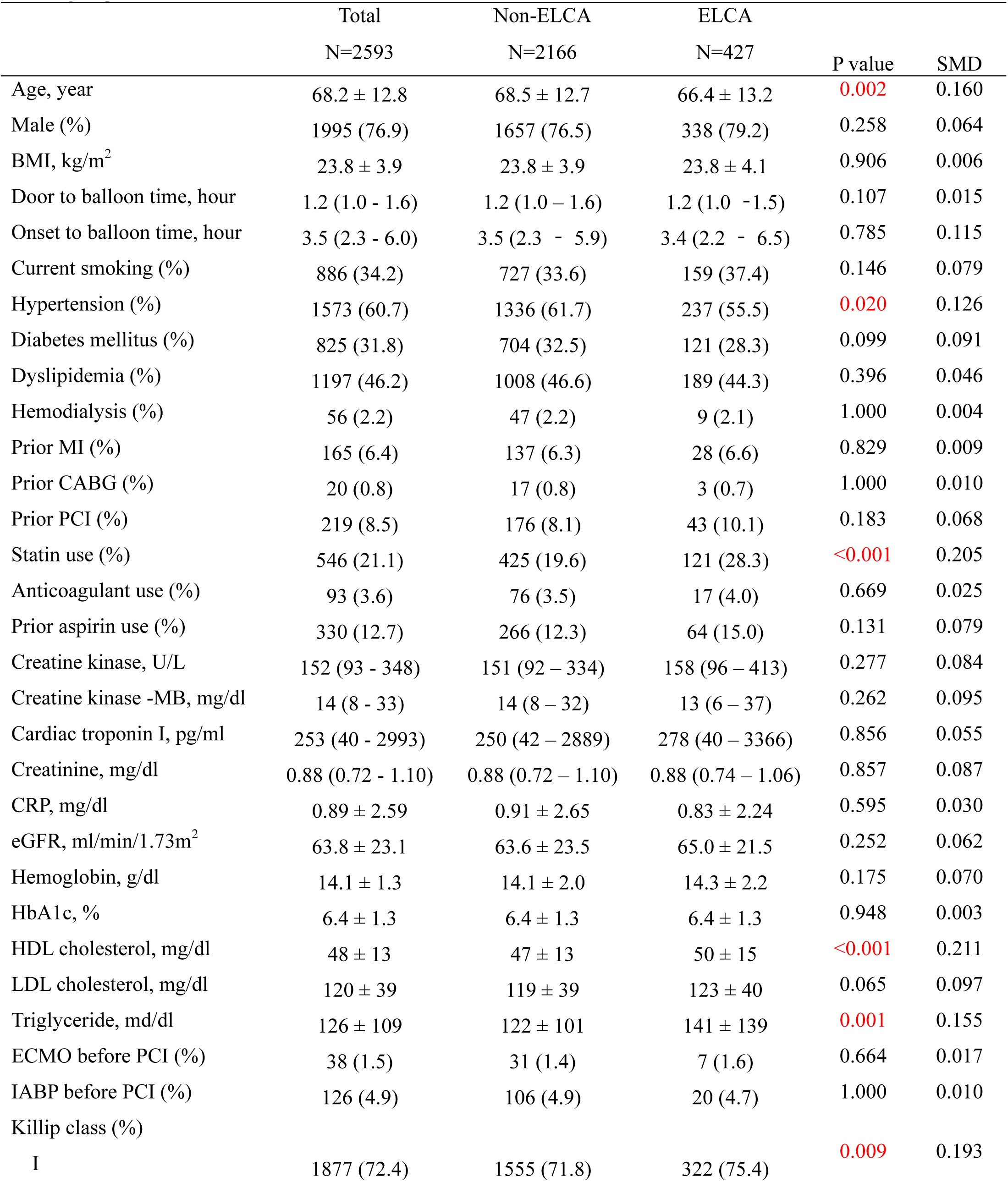
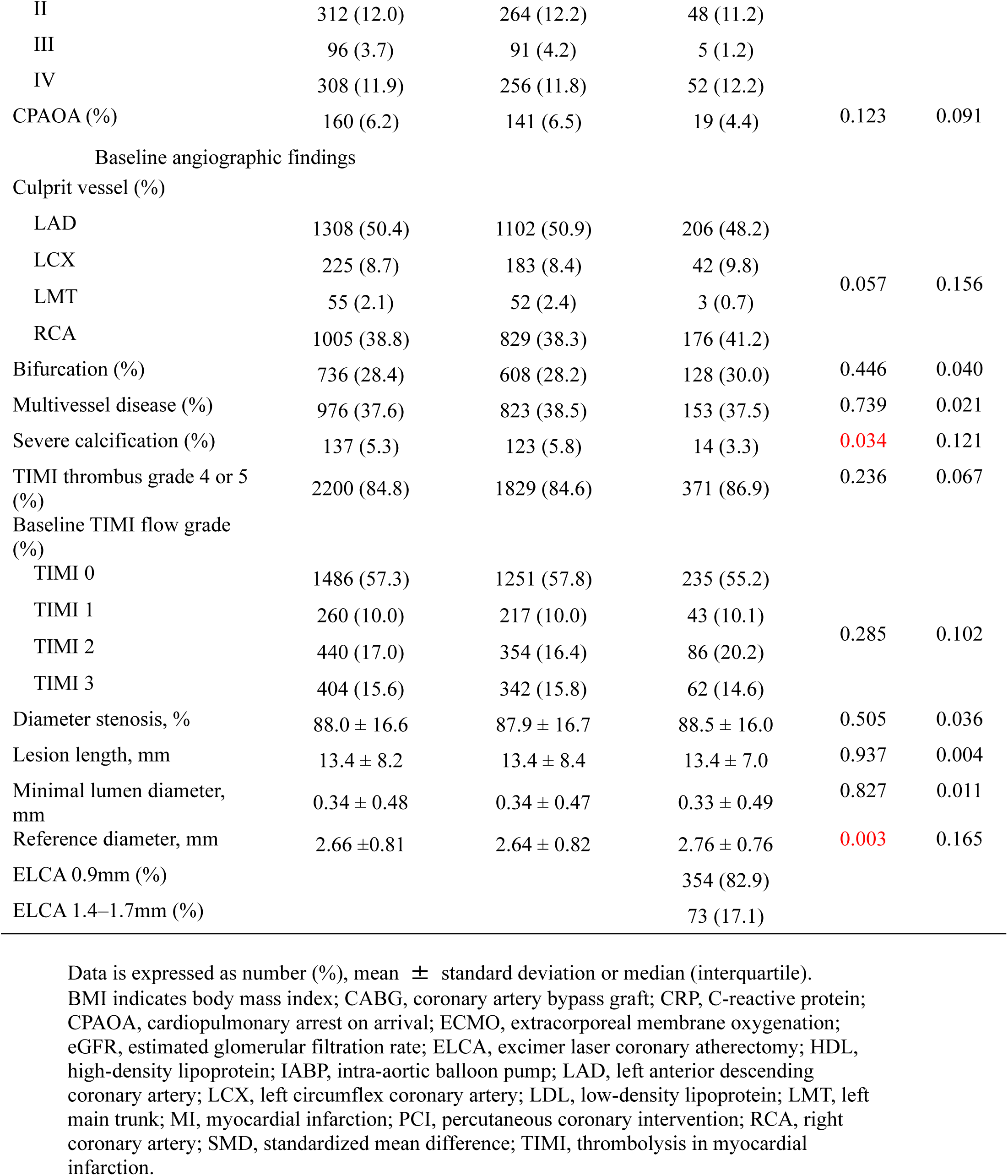
Baseline demographic and angiographic characteristics of non-ELCA and ELCA groups in the total cohort.

|  | Total<br>N=2593 | Non-ELCA<br>N=2166 | ELCA<br>N=427 | P value | SMD |
| --- | --- | --- | --- | --- | --- |
| Age, year | 68.2 ± 12.8 | 68.5 ± 12.7 | 66.4 ± 13.2 | 0.002 | 0.160 |
| Male (%) | 1995 (76.9) | 1657 (76.5) | 338 (79.2) | 0.258 | 0.064 |
| BMI, kg/m<sup>2</sup> | 23.8 ± 3.9 | 23.8 ± 3.9 | 23.8 ± 4.1 | 0.906 | 0.006 |
| Door to balloon time, hour | 1.2 (1.0 - 1.6) | 1.2 (1.0 – 1.6) | 1.2 (1.0 - 1.5) | 0.107 | 0.015 |
| Onset to balloon time, hour | 3.5 (2.3 - 6.0) | 3.5 (2.3 - 5.9) | 3.4 (2.2 - 6.5) | 0.785 | 0.115 |
| Current smoking (%) | 886 (34.2) | 727 (33.6) | 159 (37.4) | 0.146 | 0.079 |
| Hypertension (%) | 1573 (60.7) | 1336 (61.7) | 237 (55.5) | 0.020 | 0.126 |
| Diabetes mellitus (%) | 825 (31.8) | 704 (32.5) | 121 (28.3) | 0.099 | 0.091 |
| Dyslipidemia (%) | 1197 (46.2) | 1008 (46.6) | 189 (44.3) | 0.396 | 0.046 |
| Hemodialysis (%) | 56 (2.2) | 47 (2.2) | 9 (2.1) | 1.000 | 0.004 |
| Prior MI (%) | 165 (6.4) | 137 (6.3) | 28 (6.6) | 0.829 | 0.009 |
| Prior CABG (%) | 20 (0.8) | 17 (0.8) | 3 (0.7) | 1.000 | 0.010 |
| Prior PCI (%) | 219 (8.5) | 176 (8.1) | 43 (10.1) | 0.183 | 0.068 |
| Statin use (%) | 546 (21.1) | 425 (19.6) | 121 (28.3) | <0.001 | 0.205 |
| Anticoagulant use (%) | 93 (3.6) | 76 (3.5) | 17 (4.0) | 0.669 | 0.025 |
| Prior aspirin use (%) | 330 (12.7) | 266 (12.3) | 64 (15.0) | 0.131 | 0.079 |
| Creatine kinase, U/L | 152 (93 - 348) | 151 (92 – 334) | 158 (96 – 413) | 0.277 | 0.084 |
| Creatine kinase -MB, mg/dl | 14 (8 - 33) | 14 (8 – 32) | 13 (6 – 37) | 0.262 | 0.095 |
| Cardiac troponin I, pg/ml | 253 (40 - 2993) | 250 (42 – 2889) | 278 (40 – 3366) | 0.856 | 0.055 |
| Creatinine, mg/dl | 0.88 (0.72 - 1.10) | 0.88 (0.72 – 1.10) | 0.88 (0.74 – 1.06) | 0.857 | 0.087 |
| CRP, mg/dl | 0.89 ± 2.59 | 0.91 ± 2.65 | 0.83 ± 2.24 | 0.595 | 0.030 |
| eGFR, ml/min/1.73m<sup>2</sup> | 63.8 ± 23.1 | 63.6 ± 23.5 | 65.0 ± 21.5 | 0.252 | 0.062 |
| Hemoglobin, g/dl | 14.1 ± 1.3 | 14.1 ± 2.0 | 14.3 ± 2.2 | 0.175 | 0.070 |
| HbA1c, % | 6.4 ± 1.3 | 6.4 ± 1.3 | 6.4 ± 1.3 | 0.948 | 0.003 |
| HDL cholesterol, mg/dl | 48 ± 13 | 47 ± 13 | 50 ± 15 | <0.001 | 0.211 |
| LDL cholesterol, mg/dl | 120 ± 39 | 119 ± 39 | 123 ± 40 | 0.065 | 0.097 |
| Triglyceride, md/dl | 126 ± 109 | 122 ± 101 | 141 ± 139 | 0.001 | 0.155 |
| ECMO before PCI (%) | 38 (1.5) | 31 (1.4) | 7 (1.6) | 0.664 | 0.017 |
| IABP before PCI (%) | 126 (4.9) | 106 (4.9) | 20 (4.7) | 1.000 | 0.010 |
| Killip class (%) |  |  |  |  |  |
| I | 1877 (72.4) | 1555 (71.8) | 322 (75.4) | 0.009 | 0.193 |
| II | 312 (12.0) | 264 (12.2) | 48 (11.2) |  |  |
| III | 96 (3.7) | 91 (4.2) | 5 (1.2) |  |  |
| IV | 308 (11.9) | 256 (11.8) | 52 (12.2) |  |  |
| CPAOA (%) | 160 (6.2) | 141 (6.5) | 19 (4.4) | 0.123 | 0.091 |
| Baseline angiographic findings |  |  |  |  |  |
| Culprit vessel (%) |  |  |  |  |  |
| LAD | 1308 (50.4) | 1102 (50.9) | 206 (48.2) |  |  |
| LCX | 225 (8.7) | 183 (8.4) | 42 (9.8) |  |  |
| LMT | 55 (2.1) | 52 (2.4) | 3 (0.7) | 0.057 | 0.156 |
| RCA | 1005 (38.8) | 829 (38.3) | 176 (41.2) |  |  |
| Bifurcation (%) | 736 (28.4) | 608 (28.2) | 128 (30.0) | 0.446 | 0.040 |
| Multivessel disease (%) | 976 (37.6) | 823 (38.5) | 153 (37.5) | 0.739 | 0.021 |
| Severe calcification (%) | 137 (5.3) | 123 (5.8) | 14 (3.3) | 0.034 | 0.121 |
| TIMI thrombus grade 4 or 5 (%) | 2200 (84.8) | 1829 (84.6) | 371 (86.9) | 0.236 | 0.067 |
| Baseline TIMI flow grade (%) |  |  |  |  |  |
| TIMI 0 | 1486 (57.3) | 1251 (57.8) | 235 (55.2) |  |  |
| TIMI 1 | 260 (10.0) | 217 (10.0) | 43 (10.1) |  |  |
| TIMI 2 | 440 (17.0) | 354 (16.4) | 86 (20.2) | 0.285 | 0.102 |
| TIMI 3 | 404 (15.6) | 342 (15.8) | 62 (14.6) |  |  |
| Diameter stenosis, % | 88.0 ± 16.6 | 87.9 ± 16.7 | 88.5 ± 16.0 | 0.505 | 0.036 |
| Lesion length, mm | 13.4 ± 8.2 | 13.4 ± 8.4 | 13.4 ± 7.0 | 0.937 | 0.004 |
| Minimal lumen diameter, mm | 0.34 ± 0.48 | 0.34 ± 0.47 | 0.33 ± 0.49 | 0.827 | 0.011 |
| Reference diameter, mm | 2.66 ± 0.81 | 2.64 ± 0.82 | 2.76 ± 0.76 | 0.003 | 0.165 |
| ELCA 0.9mm (%) |  |  | 354 (82.9) |  |  |
| ELCA 1.4–1.7mm (%) |  |  | 73 (17.1) |  |  |

Data is expressed as number (%), mean ± standard deviation or median (interquartile). BMI indicates body mass index; CABG, coronary artery bypass graft; CRP, C-reactive protein; CPAOA, cardiopulmonary arrest on arrival; ECMO, extracorporeal membrane oxygenation; eGFR, estimated glomerular filtration rate; ELCA, excimer laser coronary atherectomy; HDL, high-density lipoprotein; IABP, intra-aortic balloon pump; LAD, left anterior descending coronary artery; LCX, left circumflex coronary artery; LDL, low-density lipoprotein; LMT, left main trunk; MI, myocardial infarction; PCI, percutaneous coronary intervention; RCA, right coronary artery; SMD, standardized mean difference; TIMI, thrombolysis in myocardial infarction.

### Trends in the use of ELCA

Of the 2593 cases, 1646 (63.5%) were enrolled from ELCA-available hospitals. In the ELCA-available hospitals, ELCA was used during primary PCI in 427 cases (25.9%), which varied in the usage rate from 0.4 to 84.5% in different facilities (Supplemental Figure 1). Of 427 patients undergoing ELCA, the maximum ELCA catheter size was 0.9 mm in 354 (82.9%) patients, while larger ELCA catheters including 1.4 mm and 1.7 mm were used in 73 (17.1%) patients. The predictors of ELCA use were determined using multiple logistic regression analyses (Table 2). ELCA tended to be used in patients with younger age, smaller body mass index, lower creatinine level, non-left main disease, greater reference lumen diameter at the initial angiogram, and TIMI grade 4 or 5 thrombus. ELCA was preferred in patients with Killip class IV disease at presentation, whereas ELCA tended to be avoided in patients with class III disease.

**Table 2.**
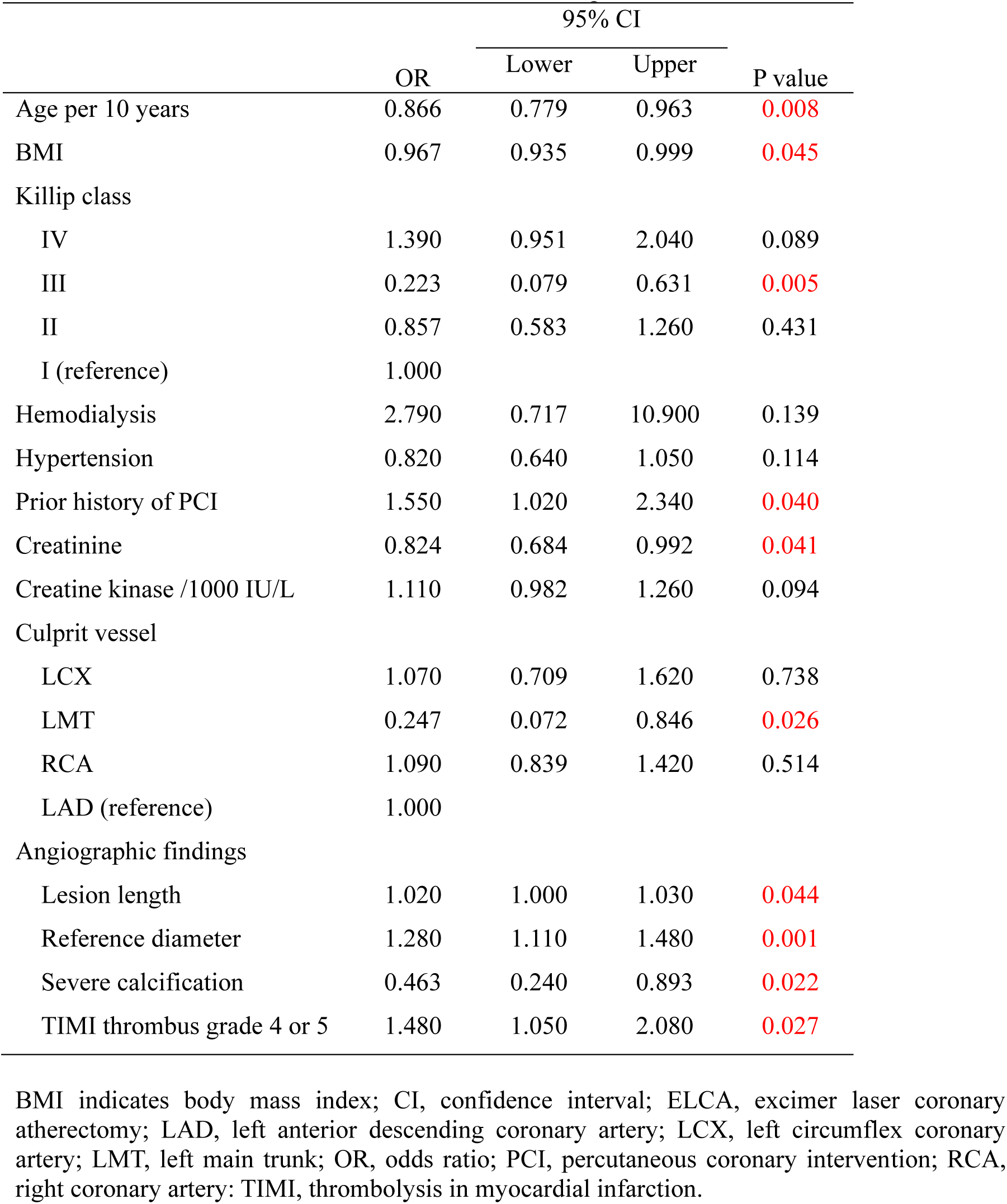
Predictors of ELCA use in ELCA-available hospitals.

### Comparison of ELCA and non-ELCA groups in the total cohort

Baseline characteristics of the ELCA and non-ELCA groups in the total cohort are shown in Table 1. The ELCA group showed younger age, less frequent hypertension, more frequent prior statin use, higher HDL cholesterol levels, higher triglyceride levels, more thrombotic lesions defined as TIMI thrombus burden class 4 or greater, and a greater angiographic reference diameter at baseline. The distribution of Killip classes differed significantly between the two groups, with less prevalent Killip III class in the ELCA group. In terms of angiographic findings, a large thrombus, defined as a TIMI grade 4 or 5 thrombus, was more frequently observed, and the reference diameter was significantly greater in the ELCA group than in the non-ELCA group. The corrected TIMI frame count was worse in the ELCA group than in the non-ELCA group, while no significant differences were observed in the QCA parameters, TIMI flow grade, or myocardial blush grade (MBG). The peak creatine kinase myocardial band (CK-MB) level was significantly higher in the ELCA group, whereas the peak cardiac troponin I level was significantly greater in the non-ELCA group. The clinical and angiographic outcomes after PCI are summarized in Table 3. During the median follow-up period of 2.2 (1.1–3.8) years, TV-MACE occurred in 355 patients (13.7%), which included 187 CVD (7.2%), 156 TVR (6.0%), and 24 TV-MI (0.9%). The primary endpoint, TV-MACE-free survival rate, did not differ between the non-ELCA and ELCA groups (Figure 2 and Figure 3). Nevertheless, when stratified by the maximum ELCA catheter size, patients treated with a large catheter, 1.4 mm or greater, showed a non-significant trend for better TV-MACE-free survival in the log-rank test (p=0.062), whereas the univariate Cox hazard model showed a significantly better outcome in the large catheter ELCA group than in the non-ELCA group [HR 0.303, 95% confidence interval (CI), 0.097–0.946; p=0.040) (Figure 3). In the multivariable Cox hazard model of TV-MACE, the culprit vessel, greater angiographic lesion length, smaller reference diameter, severe angiographic calcification, Killip class IV, higher creatine kinase (CK) level, lower triglyceride level, hemodialysis, previous history of PCI, and the requirement for extracorporeal membrane oxygenation (ECMO) or intra-aortic balloon pumping (IABP) before PCI were significant predictors of TV-MACE. The use of ELCA was not significantly associated with TV-MACE (Table 4), and the use of large-sized catheter for ELCA did not remain a significant predictor of TV-MACE in multivariable analysis (HR 0.311 [95% CI 0.077–1.264], p = 0.103) (Supplemental Table 1). CVD was significantly less frequently observed in the ELCA group than in the non-ELCA group (Table 3 and Figure 2B). However, in the Cox hazard model to predict CVD, ELCA did not remain a significant predictor (HR 0.951 [95% CI 0.529–1.712]). Instead, smaller minimal lumen diameter, severe calcification, lower TIMI thrombus grade, age, greater Killip class, higher BMI, higher creatinine, lower triglycerides, prior history of PCI, and the requirement for IABP before PCI were associated with CVD (Supplemental Table 2). The TVR-free survival rate was significantly lower in the ELCA group than in the non-ELCA group (Figure 2C), even after adjusting, in the multivariate Cox hazard model, for longer angiographic lesion length, non-left circumflex lesions, younger age, greater CK level, hemodialysis, and prior statin use (Supplemental Table 3). The rate of target vessel-related MI did not differ between the ELCA and non-ELCA groups (Figure 2D). In multivariate Cox regression analysis, younger age, lower BMI, diabetes mellitus, and in-stent thrombosis as the culprit lesion for STEMI were significantly associated with TV-MI (Supplemental Table 4).

**Figure 2.**
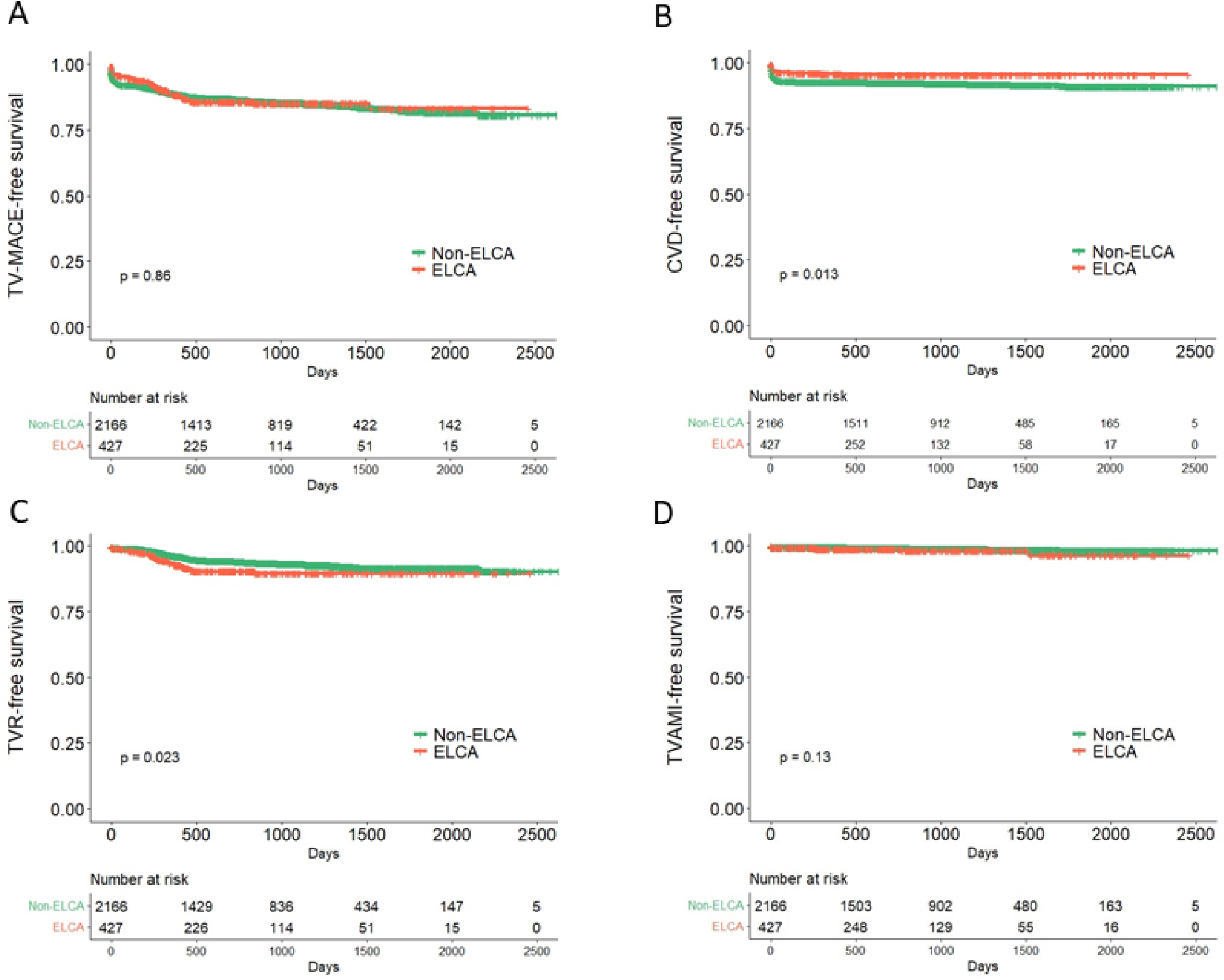
Event-free survival rate in the total cohort. Each free survival rate in ELCA group and non-ELCA group. A. Target vessel-related major adverse cardiac events B. Cardiovascular death. C. Target-vessel revascularization. D. Target-vessel-related non-fatal acute myocardial infarction. ECLA, excimer laser coronary atherectomy.

**Figure 3.**
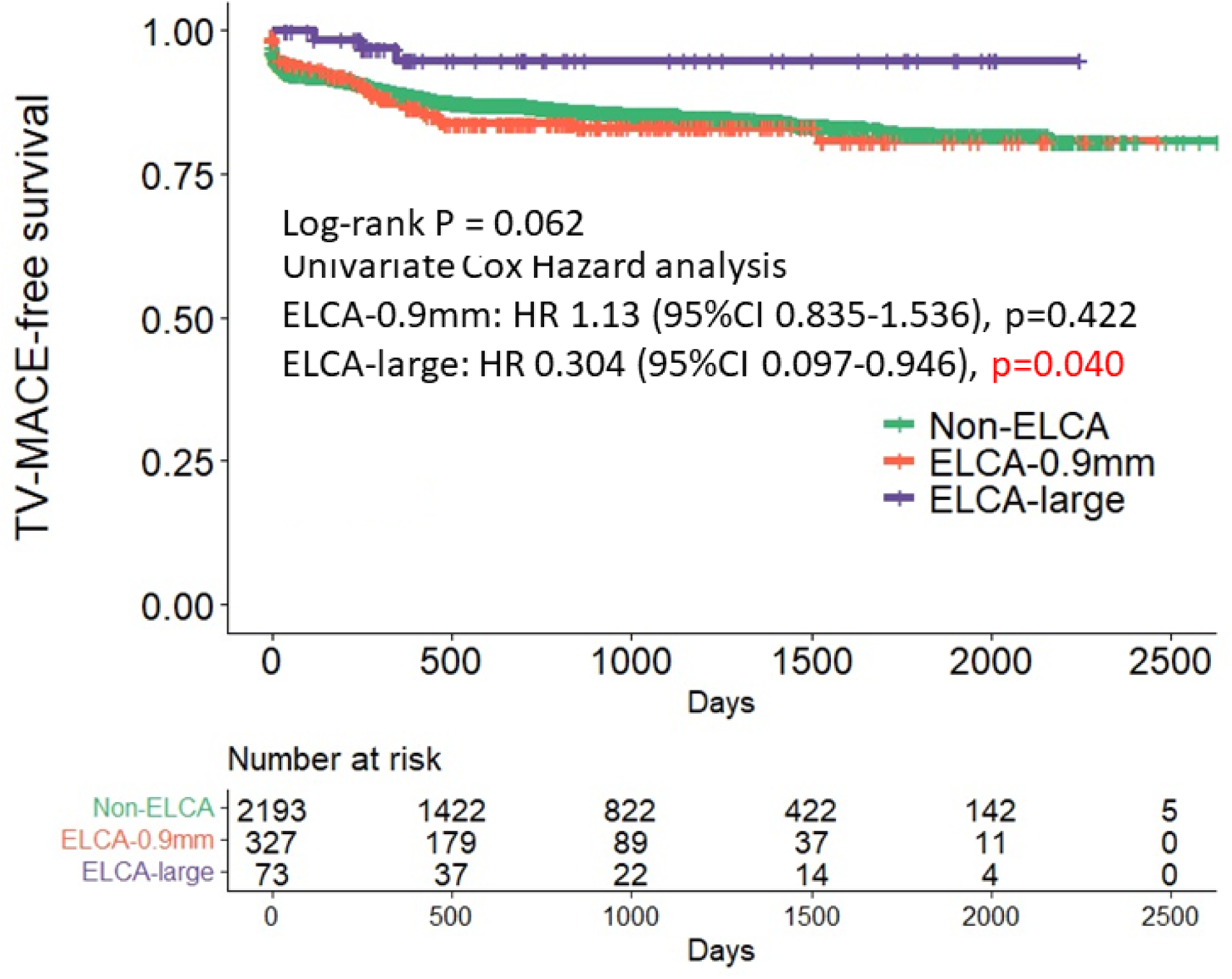
TV-MACE free survival according to ELCA size. The target vessel-related major adverse cardiac events (TV-MACE)-free survival rate was determined after stratifying by the size of the ELCA catheter. (non-ELCA, ELCA-0.9 mm, and large ELCA indicating). The large catheter ELCA group (1.4 mm or larger catheter) showed a lower risk of TV-MACE than the 0.9 mm catheter ECLA and non-ELCA groups in the univariate Cox hazard regression analysis.

**Table 3.** Clinical outcomes and angiographic findings according to the ELCA use in the total cohort.

|  | Non-ELCA | ELCA |  |  |
| --- | --- | --- | --- | --- |
|  | N=2166 | N=427 | P value | SMD |
| Peak CK level, U/l | 2182 (1040 – 4273) | 2489 (1098 – 4364) | 0.278 | 0.018 |
| Peak CK-MB level, mg/dl | 201 (91 – 393) | 242 (102 – 423) | 0.038 | 0.066 |
| Peak cTnI level, pg/dl | 36549<br>(3073 – 128772) | 29540<br>(2285 – 105903) | 0.044 | 0.159 |
| Angiographic findings at the final angiogram during primary PCI |  |  |  |  |
| Diameter stenosis, % | 18.7 ± 12.1 | 19.0 ± 12.2 | 0.677 | 0.022 |
| Minimal lumen diameter, mm | 2.6 ± 0.6 | 2.5 ± 0.6 | 0.326 | 0.052 |
| Reference diameter, mm | 3.1 ± 0.6 | 3.1 ± 0.7 | 0.439 | 0.040 |
| Corrected TIMI frame count | 25.0 (17.7 – 38.0) | 27.0 (18.0 – 41.0) | 0.015 | 0.134 |
| Slow flow (%) | 355 (16.4) | 69 (16.2) | 0.943 | 0.006 |
| TIMI flow grade |  |  |  |  |
| grade 0 | 20 (0.9) | 1 (0.2) | 0.438 | 0.100 |
| grade 1 | 38 (1.8) | 10 (2.3) |  |  |
| grade 2 | 287 (13.3) | 58 (13.6) |  |  |
| grade 3 | 1811 (84.0) | 358 (83.8) |  |  |
| Myocardial blush grade |  |  |  |  |
| grade 0 | 16 (0.7) | 6 (1.4) | 0.327 | 0.094 |
| grade 1 | 77 (2.6) | 11 (2.6) |  |  |
| grade 2 | 531 (24.7) | 99 (23.2) |  |  |
| grade 3 | 1522 (70.9) | 311 (72.8) |  |  |
| Clinical events |  |  |  |  |
| TV-MACE (%) | 301 (13.9) | 54 (12.6) | 0.538 | 0.037 |
| Cardiovascular death (%) | 169 (7.8) | 18 (4.2) | 0.008 | 0.151 |
| TVR (%) | 123 (5.7) | 33 (7.7) | 0.118 | 0.082 |
| Target vessel AMI (%) | 18 (0.8) | 6 (1.4) | 0.266 | 0.055 |
| Non-target vessel related revascularization (%) | 142 (6.6) | 36 (8.4) | 0.173 | 0.071 |
| Any nonfatal MI (%) | 41 (1.9) | 7 (1.6) | 0.846 | 0.019 |

Data is expressed as number(%), mean ± standard deviation or median (interquartile). AMI indicates acute myocardial infarction; CK, creatine kinase; ELCA, excimer laser coronary atherectomy; MI, myocardial infarction; SMD, standardized mean difference; TV-MACE, target vessel major adverse cardiac event; TIMI, thrombolysis in myocardial infarction; TVR, target vessel revascularization.

**Table 4.** Predictors of target vessel related major adverse cardiac events in the total cohort.

|  |  | 95% CI |  |  |
| --- | --- | --- | --- | --- |
|  | HR | Lower | Upper | P value |
| Culprit vessel |  |  |  |  |
| LCX | 0.403 | 0.211 | 0.769 | 0.006 |
| LMT | 1.890 | 1.005 | 3.554 | 0.048 |
| RCA | 1.009 | 0.772 | 1.320 | 0.946 |
| LAD (reference) | 1 |  |  |  |
| Lesion length at the baseline | 1.014 | 1.001 | 1.026 | 0.033 |
| Reference diameter baseline | 0.833 | 0.702 | 0.989 | 0.037 |
| Angiographic severe calcification | 1.925 | 1.268 | 2.922 | 0.002 |
| Killip class |  |  |  |  |
| IV | 3.159 | 2.251 | 4.434 | <0.001 |
| III | 1.405 | 0.769 | 2.568 | 0.268 |
| II | 0.992 | 0.660 | 1.490 | 0.967 |
| I (reference) | 1 |  |  |  |
| ELCA use | 1.265 | 0.910 | 1.757 | 0.161 |
| Creatine kinase per1000 IU/L | 1.114 | 1.025 | 1.210 | 0.011 |
| Triglyceride | 0.997 | 0.996 | 0.999 | 0.004 |
| Hemodialysis | 3.701 | 2.220 | 6.172 | <0.001 |
| Prior PCI | 2.099 | 1.480 | 2.976 | <0.001 |
| ECMO before PCI | 2.373 | 1.264 | 4.455 | 0.007 |
| IABP before PCI | 1.899 | 1.216 | 2.968 | 0.005 |

CI indicates confidence interval; ECMO, extracorporeal membrane oxygenation; ELCA, excimer laser coronary atherectomy; HR, hazard ratio; IABP, intra-aortic balloon pump; LAD, left anterior descending artery; LCX, left circumflex coronary artery; LMT, left main trunk; PCI, percutaneous coronary intervention; RCA, right coronary artery.

### PS-matched ELCA and non-ELCA groups

The PS was derived from clinical features including age, sex, onset-to-balloon time, door-to-balloon time, Killip class at presentation, angiographic findings including culprit vessel location, TIMI flow grade, TIMI thrombus grade, minimal lumen diameter, reference diameter, lesion length, and in-stent lesion; laboratory data including CK level at presentation, creatinine, HbA1c, LDL cholesterol, and HDL cholesterol levels; demographic history including current smoking, diabetes, hypertension, dyslipidemia, prior statin use, and prior aspirin use; and procedural conditions including ECMO and IABP use before PCI. A total of 736 patients (368 matched pairs) were selected for PSM analysis. The baseline demographic and angiographic data of the PSM cohort were well balanced between the non-ELCA and ELCA groups (Supplemental Table 5). The Kaplan–Meier curves for adverse events are summarized in Figure 4. There were no significant differences in the event-free survival rates for TV-related MACE, CVD, TVR, or TV-MI in the log-rank test. Table 5 compares the clinical and angiographic secondary endpoints of the two PSM groups. Peak CK and CK-MB levels were significantly higher in the ELCA group than in the non-ELCA group, which was consistent with the results of the total cohort, whereas no significant difference was observed in peak cardiac troponin-I levels. There were no significant differences in angiographic results except for a slight deterioration of the corrected TIMI frame count in the ELCA group.

**Figure 4.**
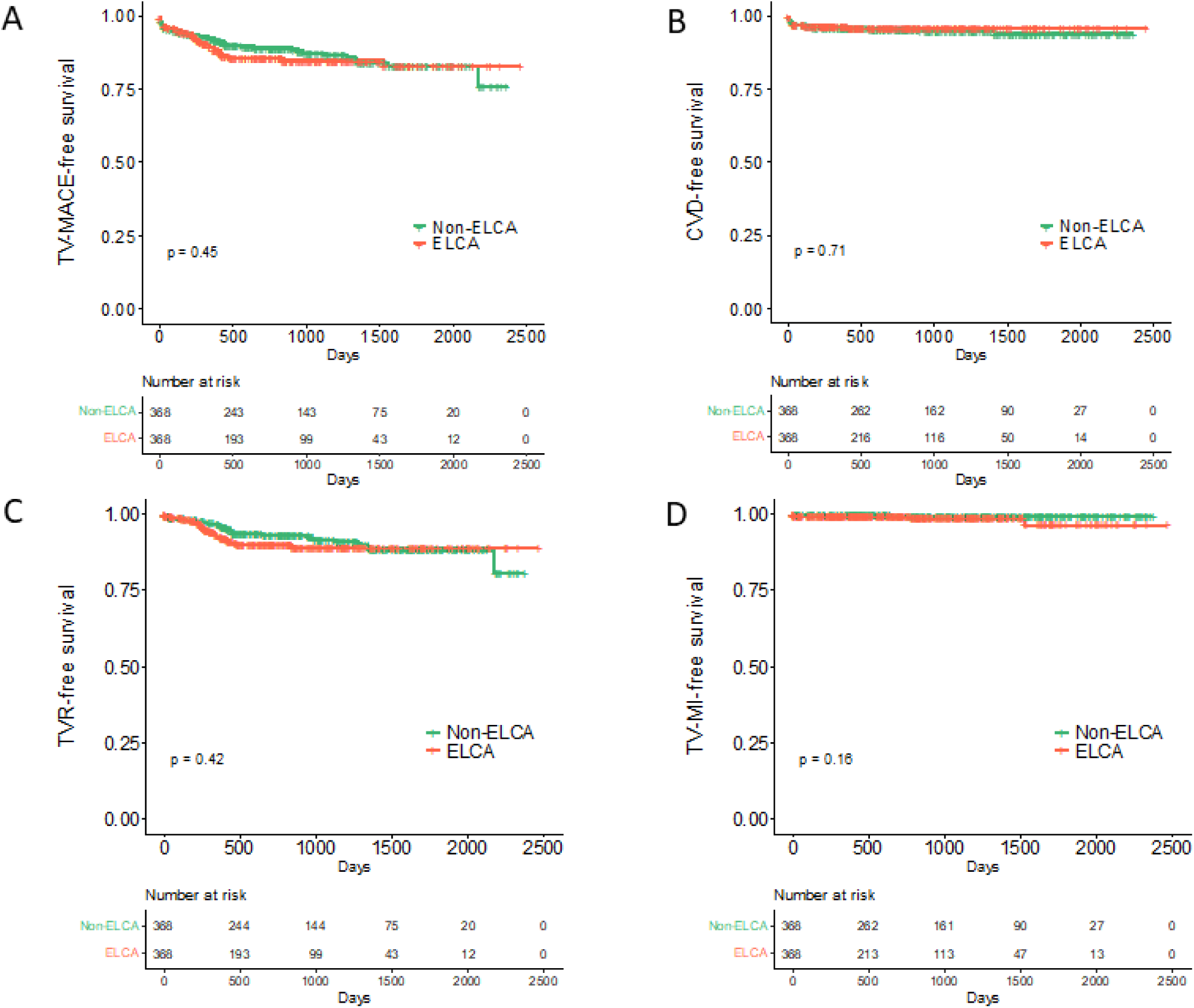
Event-free survival rate in the propensity score matched cohort. Each free survival rate in ELCA group and non-ELCA group. A. Target vessel-related major adverse cardiac events B. Cardiovascular death. C. Target-vessel revascularization. D. Target-vessel-related non-fatal acute myocardial infarction. ECLA, excimer laser coronary atherectomy.

**Table 5.** Clinical outcomes and angiographic findings according to the ELCA use in the matched cohort.

|  | Non-ELCA | ELCA |  |  |
| --- | --- | --- | --- | --- |
|  | N=368 | N=368 | P value | SMD |
| Peak CK level, U/l | 2048 (926–3895) | 2449 (1072–4415) | 0.044 | 0.107 |
| Peak CK-MB level, md/dl | 193 (83–361) | 241 (95–410) | 0.011 | 0.162 |
| Peak cTnI level, pg/ml | 25288<br>(1037–106252) | 31200<br>(2025–105903) | 0.776 | 0.082 |
| Angiographic findings at the final angiogram during primary PCI |  |  |  |  |
| Diameter stenosis, % | 17.8 ± 11.6 | 18.9 ± 11.4 | 0.188 | 0.097 |
| Minimal lumen diameter, mm | 2.6 ± 0.6 | 2.5 ± 0.6 | 0.524 | 0.047 |
| Reference diameter, mm | 3.1 ± 0.6 | 3.1 ± 0.7 | 0.613 | 0.037 |
| Corrected TIMI frame count | 24.0 (16.0–34.0) | 26.0 (18.0–39.0) | 0.012 | 0.223 |
| Slow flow (%) | 40 (10.9) | 58 (15.8) | 0.065 | 0.144 |
| TIMI flow grade |  |  |  |  |
| grade 0 | 1(0.3) | 1(0.3) | 0.180 | 0.154 |
| grade 1 | 5(1.4) | 7(1.9) |  |  |
| grade 2 | 33(9.0) | 50(13.6) |  |  |
| grade 3 | 328(89.4) | 310(84.2) |  |  |
| Myocardial blush grade |  |  | 0.547 | 0.109 |
| grade 0 | 1 (0.3) | 3 (0.8) |  |  |
| grade 1 | 10 (2.7) | 9 (2.4) |  |  |
| grade 2 | 74 (20.2) | 86 (23.4) |  |  |
| grade 3 | 281 (76.8) | 270 (73.4) |  |  |
| Clinical events |  |  |  |  |
| TV-MACE (%) | 45(12.2) | 46(12.5) | 1.000 | 0.008 |
| Cardiovascular death (%) | 17(4.6) | 14(3.8) | 0.714 | 0.041 |
| TVR (%) | 29(7.9) | 30(8.2) | 1.000 | 0.010 |
| Target vessel AMI (%) | 2(0.5) | 5(1.4) | 0.451 | 0.084 |
| Non-target vessel related revascularization (%) | 27(7.3) | 31(8.4) | 0.682 | 0.040 |
| Any nonfatal MI | 4(1.1) | 6(1.6) | 0.752 | 0.047 |

Data is expressed as number(%) or average standard ± deviation or median (interquartile). Abbreviation is indicated in Table 3.

## Discussion

This study investigated the efficacy of ELCA for culprit lesions in 2593 patients with STEMI, including 427 treated with ELCA. The major findings of the study were: 1) during a median follow-up period of 2.2 years, the ELCA group did not show a significant advantage over the non-ELCA group in terms of TV-MACE, which was defined as a composite endpoint of CVD, TVR, and TV-MI; 2) the ELCA group showed higher TVR rates and lower CVD rates in the total cohort, however, these differences were not significant after adjusting for variables in the Cox hazard model; 3) stratification of the ELCA group by ELCA catheter size showed a trend toward an improved TV-MACE-free rate with larger catheter size (1.4 mm to 1.7 mm), compared to both the 0.9 mm catheter ELCA group and the non-ELCA group; and 4) in the PSM cohort, there were no significant differences in clinical events between the ELCA and non-ELCA groups, although the ELCA group exhibited higher CK and CK-MB levels and worse corrected TIMI frame counts than the non-ELCA group.

### The use of ELCA for thrombotic lesions

Unlike the early forms of laser atherectomy including argon laser^15,16^, the xenon chloride excimer laser emits energy in the ultraviolet range (308 nm) in extremely short pulses. This unique characteristic allows for procedural safety due to its limited absorption depth (<100 µm) and reduces the risk of collateral tissue damage. ELCA was approved by the United States Food and Drug Administration in 1992. ELCA ablates arterial tissue via three major mechanisms: photochemical, photothermal, and photokinetic effects, which lead to the fracture of molecular bonds, tissue vaporization, and clearance of byproducts within the target tissue.^17^ These mechanisms are particularly effective in modifying plaques that are challenging for other devices, such as lesions that are balloon-undilatable or uncrossable. Consequently, ELCA received a Class IIb recommendation in the U.S. guidelines for plaque modification, ^18^ especially for fibrotic or heavily calcified lesions, ^19,20^ in addition to rotational or orbital atherectomy. Additionally, thrombi are potential targets for ELCA in angioplasty. A xenon chloride laser is absorbed by the thrombus materials, which induces selective thrombolysis, inhibits the local aggregation of platelets,^21^ and ablates the atherosclerotic components underneath. Based on the theoretical background of the effectiveness of ELCA for thrombectomy, it has been used as an adjunctive tool to reduce thrombus burden during PCI without concrete evidence.^6^ In the present study, physicians in ELCA-available facilities tended to use ELCA in patients with a greater reference vessel diameter and TIMI thrombus grade (Table 1).

### Clinical outcomes after ELCA-assisted primary PCI

Given the mechanisms of thrombus formation and the large volume of plaque, ACS may theoretically receive the maximal benefit of ELCA. Topaz et al. reported the use of ELCA in 59 patients with ACS and demonstrated its safety.^7^ The Cohort of Acute Revascularization in Myocardial Infarction with Excimer Laser (CAMEL) trial was the first prospective study to investigate the efficacy of ELCA for the treatment of culprit AMI lesions.^22^ This single-armed, multicenter trial used ELCA during primary PCI in patients with AMI within 24 h from onset, and reported reduced angiographic thrombus and improved coronary flow after ELCA use, although there was no non-ELCA control group. Shishikura et al. compared MBG and ST-segment resolution between 50 patients with ACS treated with ELCA and 48 age- and sex-matched patients with ACS not receiving ELCA, and reported that ELCA treatment led to better MBG and ST-segment resolution.^8^ However, the clinical benefit of ELCA in primary PCI remains controversial, and there is a paucity of evidence regarding the reduced incidence of major adverse events after ELCA compared to standard procedures without ELCA. In fact, in some reports, the effectiveness of ELCA was reduced in lesions with angiographic thrombus.^4^ Furthermore, the only randomized controlled trial, consisting of 27 patients, failed to demonstrate clinical benefits of ELCA in patients with AMI.^11^ The present study sought to identify the clinical outcomes by comparing patients with STEMI who underwent primary PCI with or without ELCA. We found that, in the total cohort of our registry data, the rate of target vessel-related adverse events was similar between the ELCA and non-ELCA groups. This finding was consistent with the results of the multivariable Cox regression and PSM analysis. In terms of cardiovascular death, the total cohort showed a lower event rate in the ELCA group than that in the non-ELCA group, which may be in line with previous single-center studies that showed preferable revascularization after ELCA. However, the ELCA group was significantly younger, and the patients’ age was a strong predictor of CVD. After adjustment by PSM, ELCA did not remain a significant factor for CVD. Thus, contrary to previous findings, the present study failed to demonstrate the benefits of ELCA in terms of the clinical endpoints.

### Angiographic findings and cardiac enzymes after ELCA-assisted primary PCI

Several studies have reported the potential efficacy of ELCA using surrogate markers of improved revascularization, such as MBG, TIMI flow grade, peak cardiac enzyme levels, and myocardial salvage assessed using scintigraphy.^8,9^ This study revealed worse post-PCI coronary flow and higher CK and CK-MB levels in the ELCA group than in the non-ELCA group in the PSM cohort, contradicting previous findings. There are several possible reasons for this discrepancy. The first is an unadjustable selection bias, particularly in single-center retrospective studies. Previous studies were conducted retrospectively in hospitals where ELCA was available, inherently introducing an unavoidable selection bias even after multivariate analysis or PSM. In other words, physicians may have tended to select more suitable patients for ELCA, a factor that cannot be fully accounted for in a single-center cohort. Therefore, the current study, which collected data not only from hospitals where ELCA was available for primary PCI but also from hospitals where it was not available, may offer a more equitable basis for comparing outcomes between the ELCA and non-ELCA groups. Another potential reason is the size of the catheter. In the current study, ELCA was employed in 427 lesions, with the 0.9 mm ELCA catheter being most used, and larger catheters were utilized in only 73 (17.1%) patients. Although a small-sample single-center study showed comparable clinical outcomes between 0.9 mm and larger ELCA catheters,^23^ the majority of previous studies demonstrating favorable outcomes with ELCA treatment predominantly used catheters of 1.4 mm or larger.^8,22^ Therefore, the difference in catheter size may have resulted in the lesser ELCA benefits in the current cohort. When we separately analyzed the clinical outcomes of patients who underwent ELCA with a large catheter, they showed a statistically borderline trend toward a better event-free rate than the non-ELCA group (Figure 3).

### Future perspective

The efficacy of ELCA in primary PCI for STEMI has not been conclusively proven in a prospective manner, and there are no recommendations in the current guidelines. Although smaller observational studies and case reports have suggested the efficacy of ELCA during primary PCI,^20,24-26^ systematic proof of its clinical benefits has not been reported. The present study, which included a substantial number of primary PCI cases using ELCA, failed to show a clinical benefit of ELCA, suggesting that the routine use of ELCA might not be effective for primary PCI. Nevertheless, there was a clue to the potential benefit of ELCA when a large ELCA catheter (1.4 mm or larger) was used, although this observation did not reach statistical significance in the multivariable analyses. The ongoing randomized controlled trial (NCT03950310) comparing myocardial damage after primary PCI for anterior STEMI may offer new insights. Prospective studies showing improved clinical outcomes, including event rate, after ELCA in patients with STEMI are needed to support the clinical use of ELCA.

### Limitations

The present study had several limitations. This was a retrospective analysis of cohort data, although it involved a large STEMI cohort, which may have unadjustable selection bias for ELCA use and other therapeutic strategies, even after multivariable analysis or PSM. All clinical and procedural decisions were made at the discretion of the physicians, which may have led to bias in the results. Laboratory data were examined at local participating sites, therefore, measurement errors may have occurred in different facilities.

### Conclusions

In a retrospective analysis of a relatively large cohort of patients with STEMI undergoing primary PCI, those treated with ELCA failed to show a better target vessel-related adverse event rate than those treated without ELCA. The use of a large diameter ELCA catheter showed a nonsignificant trend toward better outcomes, which may warrant investigation in a prospective study.

## Data Availability

Please contact the corresponding author for the dataset.

## Nonstandard abbreviations and acronyms

ACS: acute coronary syndrome
CK-MB: creatine kinase myocardial band
CVD: cardiovascular death
ECMO: extracorporeal membrane oxygenation
ELCA: excimer laser coronary atherectomy
IABP: intra-aortic balloon pumping
MBG: myocardial blush grade
PCI: percutaneous coronary intervention
PS: propensity score
PSM: propensity score matching
QCA: quantitative coronary angiography
STEMI: ST-segment elevation myocardial infarction
TV-MI: target vessel non-fatal myocardial infarction
TV-MACE: target vessel-related major adverse cardiac events
TVR: target vessel revascularization

## Acknowledgements

We appreciated all investigators who collaborated with and collected data with the STELA registry.

## Figure legends

**Supplemental Figure 1. Number of excimer laser coronary atherectomies performed in participating sites**

The vertical axis represents the number of cases and the horizontal axis represents each participating institution. ELCA, excimer laser coronary atherectomy.

